# Measuring the fitted filtration efficiency of cloth masks, medical masks and respirators

**DOI:** 10.1101/2024.03.17.24304429

**Authors:** Amanda A Tomkins, Gurleen Dulai, Ranmeet Dulai, Sarah Rassenberg, Darren Lawless, Scott Laengert, Rebecca S Rudman, Shiblul Hasan, Charles-Francois de Lannoy, Ken G Drouillard, Catherine M Clase

## Abstract

**Importance:** Masks reduce transmission of SARS-CoV2 and other respiratory pathogens. Comparative studies of the fitted filtration efficiency of different types of masks of are few.

**Objective:** To describe the fitted filtration efficiency against small aerosols (0.02 – 1 µm) of medical and non-medical masks and respirators when worn, and how this is affected by user modifications (hacks) and by overmasking with a cloth mask.

**Design:** We tested a 2-layer woven-cotton cloth mask of a consensus design, ASTM-certified level 1 and level 3 masks, a non-certified mask, KF94s, KN95s, an N95 and a CaN99.

**Setting:** Closed rooms with ambient particles supplemented by salt particles.

**Participants:** 12 total participants; 21 – 55 years, 68% female, 77% white, NIOSH 1 to 10.

**Main Outcome and Measure:** Using standard methods and a PortaCount 8038, we counted 0.02–1µm particles inside and outside masks and respirators, expressing results as the percentage filtered by each mask. We also studied level 1 and level 3 masks with earguards, scrub caps, the knot-and-tuck method, and the effects of braces or overmasking with a cloth mask.

**Results:** Filtration efficiency for the cloth mask was 47-55%, for level 1 masks 52-60%, for level 3 masks 60-77%. A non-certified KN95 look-alike, two KF94s, and three KN95s filtered 57-77%, and the N95 and CaN99 97-98% without fit testing. External braces and overmasking with a well-fitting cloth mask increased filtration, but earguards, scrub caps, and the knot-and-tuck method did not.

**Limitations:** Limited number of masks of each type sampled; no adjustment for multiple comparisons.

**Conclusions and Relevance:** Well-fitting 2-layer cotton masks filter in the same range as level 1 masks when worn: around 50%. Level 3 masks and KN95s/KF94s filter around 70%. External braces or overmasking with a cloth-mask-on-ties produced filtration around 90%. Only N95s and CaN99s, both of which have overhead elastic, performed close to the occupational health and safety standards for fit tested PPE (>99%), filtering at 97-99%, without fit testing. These findings inform public health messaging about relative protection from aerosols from different mask types and increase understanding of findings of studies of implementation of masks and respirators.

**Key Points:** Question: How well do medical and non-medical masks filter aerosols when worn?

Findings: Well-fitting 2-layer cotton masks, and level 1 medical masks were similar, both filtering around 50% of aerosols. Level 3 masks and KN95/KF94s were similar, filtering around 70%. N95s and CaN99s, without formal fit testing, filtered 97-98%.

Meaning: Level 1 medical masks were not better than the well-fitting 2-layer cotton masks we tested. KN95/KF94s are not as efficient, when worn, as N95s and CaN99s. Overmasking and the use of external braces improve filtration: these are potentially useful strategies when N95s are not available.

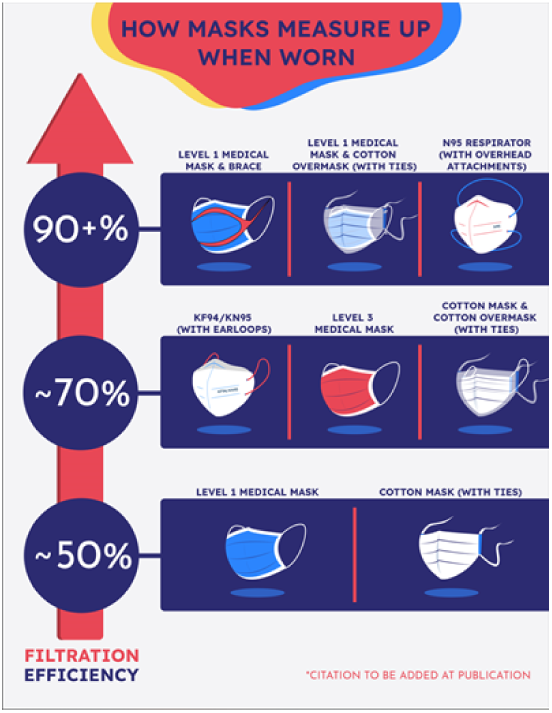

## Introduction

The global shortage of personal protective equipment (PPE) during the COVID-19 pandemic motivated inquiry into the effectiveness of public use of cloth masks.^1–16^ Current WHO guidance recommends wearing a mask as a normal part of being around other people.^17^ With increased manufacturing of masks and PPE, a variety of masks and respirators are available, including home-sewn and commercial cloth masks, certified medical masks and non-certified disposable masks, KN95/KF94s and N95/CaN99s.

Aerosol transmission of COVID-19 is now widely accepted.^18–23^ Systematic evaluation of the filtration of aerosols, from the perspective of the wearer, across the whole range of masks and non-fit-tested respirators is highly relevant to public health advice. Descriptions of cloth masks as effective only against droplets^24^ or as source control reflects and perpetuates public misunderstanding. We extend knowledge in this area by using an adequately-powered sample and a variety of masks, and by fully characterizing participants, masks, materials and modifications.

## Methods

### Population and Sampling strategy

Between May 2020 and Feb 2022, we recruited adults (≥16y), purposively sampling people of non-European ancestry, excluding people with respiratory diseases or allergic to latex. Investigators were included as participants. The Hamilton Integrated Research Ethics Board approved the study, safety procedures were developed, and participants gave informed consent.

### Procedures

#### Facial Measurement

We measured participants’ faces^25^ according to the US National Institute of Occupational Health and Safety (NIOSH) Bivariate Panel (supplementary materials, p1-11).^26^

#### Masks

We studied level 1 masks (Polar Bear and O2) and level 3 masks (Halyard and Primed), certified to the standards of ASTM International^27–29^ and donated by Hamilton Health Sciences, Hamilton, ON, Canada; these are worn on elastic earloops (supplementary figure 1). A two-layer pleated mask of woven cotton without a nosewire, the Essex mask, designed by a consensus panel for the Windsor-Essex Sewing Force (WESF) was studied both on ¼ inch elastic earloops and on overhead cloth ties (https://maskevidence.org/patternsinstruction).^30, 31^ We purposively sampled non-certified masks, KN95s, KF94s (supplementary materials, p18). We purchased 3M N95 Aura 1870+ respirators from Steripro Canada PPE Store and Vitacore CaN99 respirators from Vitacore Industries Inc, Burnaby, BC. We did not formally fit-test the respirators using occupational health and safety protocols.

#### Ambient Particle Count

We used a TSI 8026 Particle Generator (TSI, Shoreview, MN), placed at least 2m from the measuring device and participant. This aerosolises dissolved sodium chloride, producing a polydisperse aerosol that dries to solid sodium chloride particles with a diameter in the range of 0.02 μm to 0.60 μm, supplementing naturally-occurring particles in the room. We proceeded with testing if the total particle count was between 2000 and 20,000. If the ambient particle count at the end of a test was not ±30% of the count at the start, we retested.

#### Filtration efficiency testing

We fitted each mask with a sampling probe (a short aluminium tube with a flange) using a TSI model 8025-N95 Fit Test Probe Kit (TSI, Shoreview, MN). For cloth masks, if we could not penetrate the mask with the probe kit, we first used a scalpel to create a mask puncture of 1 mm. We placed the sampling port at the center of the mask; for masks with a central seam, we placed the port in the lateral flat surface. The probe attaches to a plastic tube that samples air from inside the mask. A second tube is suspended by a lanyard around the participant’s neck so that its opening is at chest height. Air sampled through this second tube reflects ambient particle counts (supplementary materials, rationale, p19-20; supplementary figure 2).

Both tubes were connected to a PortaCount 8038 (TSI Inc, Shoreview, MN).^32^ We used CSA-Z94.4-2002 protocol in all-particles mode (ie, with the N95 classifier off) to detect particles in the range 0.02-1 µm.^32, 33^ Participants donned masks and adjusted them to achieve the best subjective fit. Participants rated each mask or combination for subjective leak, and for glasses fog (using their own or safety glasses). After removing eyewear, participants conducted each of the following exercises, for 37 seconds each, according to protocol: normal breathing, deep breathing, head turning, head nodding, talking, bending and a second period of normal breathing. Finally, participants rated each mask or combination for discomfort, using Likert scales with anchors developed for this study (supplementary material, p 22). Participants also reported specific issues with masks and provided subjective free-form comments.

Fit factor for each exercise is a dimensionless number, defined as the ratio of ambient to within-mask particle count, and calculated as the geometric mean of the fit factor across all exercises for one participant. We transformed individual fit factor value to fitted filtration efficiency, as a percentage, using:^2^

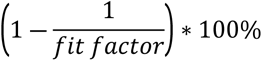

Fitted filtration efficiency describes the percentage of particles filtered by the mask. A fit factor of 4, reflecting four times the particles outside compared with inside, has a fitted filtration efficiency of 75%.

#### Community and participant involvement

The Cloth Mask Knowledge Exchange is a stakeholder group formed by the Centre of Excellence in Protective Equipment and Materials at McMaster in 2020 (supplementary materials, p23-24). This group gave input throughout the design and conduct of the experiments.

#### Statistical methods

We used SYSTAT v13, Inpixon, Palo Alto, CA for statistical analyses. We analyzed the transformed variable filtration efficiency. We used parametric tests (ANOVA with post hoc Tukey’s honestly significant difference) when data met normality assumptions by Lilliefors test; and non-parametric tests (Kruskal-Wallis with post-hoc Conover-Inman pairwise comparisons) when they did not, estimating mean and 95% confidence intervals or median and interquartile range, respectively, and regarding p<0.05 as statistically significant.

#### Studies

##### Mask types

On 4 participants, we studied the filtration efficiency of cloth masks on earloops and on overhead ties, level 1 and level 3 certified medical masks (two of each, on earloops), a KN95, two KN94s, a non-certified KN-95-lookalike mask, an N95 and a CaN99 (supplementary figure 1).

##### Mask hacks

On 10 participants, we studied the filtration efficiency of ASTM level 1 (Polar Bear) and level 3 (Halyard) masks, worn as intended, and with minor modifications or hacks (supplementary figure 1).^5, 34–36^ Three participants had short beards.^37, 38^

##### Overmasking

On 6 participants, in triplicate, we studied overmasking, using an Essex mask on earloops, a level 1 (Polar Bear) and a level 3 (Halyard) certified mask as the base mask, and overmasking with either an Essex mask on earloops or an Essex mask with overhead ties (supplementary figure 1).

#### Power calculation

In preliminary data, mean filtration efficiency was 50%, standard deviation 10%, for level 1 masks and cloth masks. For the study of mask types, the aim was to show the range of absolute filtration for each; using 4 participants gives 95% confidence intervals of 50-70% around a mean of 60% and 80-100% around a mean of 90%. For mask hacks and overmasking, we were interested in detecting differences of 10%; at alpha 0.05 and 80% power, we calculated that 6 people were needed to test each mask.^39^ To improve power and generalizability, we recruited 10 participants to the study of mask hacks, and to improve power, we performed measurements in triplicate on 6 participants in the overmasking study.

## Results

The 12 participants were 21 – 55 years, 58% female, 25% non-European, NIOSH 1 to 10 (table and supplementary table 1).

**Table.** Summary demographic and anthropometric data on participants.

| Study | Mask Types | Mask Hacks | Overmasking |
| --- | --- | --- | --- |
| Number of Participants | 4 | 10 | 6 |
| Number of Replicates | 1 | 1 | 3 |
| Age, y (mean, SD) | 30, 17 | 35, 17 | 38, 18 |
| Women/Men | 4/0 | 5/5 | 5/1 |
| Facial hair (n, %) | 0 | 3 | 0 |
| Self-identified Ethnicity (n, %) |  |  |  |
| European | 2 | 9 | 4 |
| West Central Asian and Middle Eastern | 2 | 1 | 2 |
| Height, m (mean, SD) | 1.67, 0.08 | 1.76, 0.08 | 1.67, 0.08 |
| Weight, kg (mean, SD) | 63, 12 | 77, 14 | 63, 16 |
| Bizygomatic distance, calipers, cm (mean, SD) | 12.3, 0.9 | 13.7, 1.1 | 12.3, 1.1 |
| Menton-sellion distance, calipers, cm (mean, SD) | 11.0, 0.8 | 11.5, 1.3 | 11.0, 1.0 |
| Bizygomatic distance, cord, cm (mean, SD) | 15.7, 1.1 | 15.2, 2.3 | 15.7, 2.2 |
| Menton-sellion distance, cord, cm (mean, SD) | 14.5, 2.5 | 15.5, 2.2 | 14.5, 2.0 |
| NIOSH Bivariate Panel Size (n, %) |  |  |  |
| 1 (small) | 1 | 2 | 1 |
| 2-4 (medium) | 1 | 2 | 2 |
| 5-7, 9 (large) | *2 | 4 | *3 |
| 8, 10 (X-large) | 0 | 2 | 0 |

SD standard deviation. NIOSH related to small, medium, large, X-large sizing according to ASTM 3502:22a<sup>60</sup>
\* Includes 2 participants with face width narrower than smallest face width on the NIOSH panel, classified in the nearest cell at NIOSH 6.
We studied participants with facial hair only in the mask hacks study. Facial hair was classified according to CDC NIOSH<sup>37</sup>

### Mask types

Tested on 4 participants, the 2-layer cotton pleated Essex mask resulted in a mean fitted filtration efficiency of 47% when worn on earloops and 54% on overhead ties (p >0.05; figure 1). These were comparable with results for the certified level 1 masks, at 52 and 56%, and one of the level 3 masks at 60% (p >0.05), but a second level 3 mask filtered at 75% (p <0.05). The KF94 and KN95s, and the KN95-lookalike mask filtered between 57% and 77%. The certified respirators (not fit-tested) filtered at 97-98%. The lowest value for an individual for either respirator was 95%.

**Figure 1.**
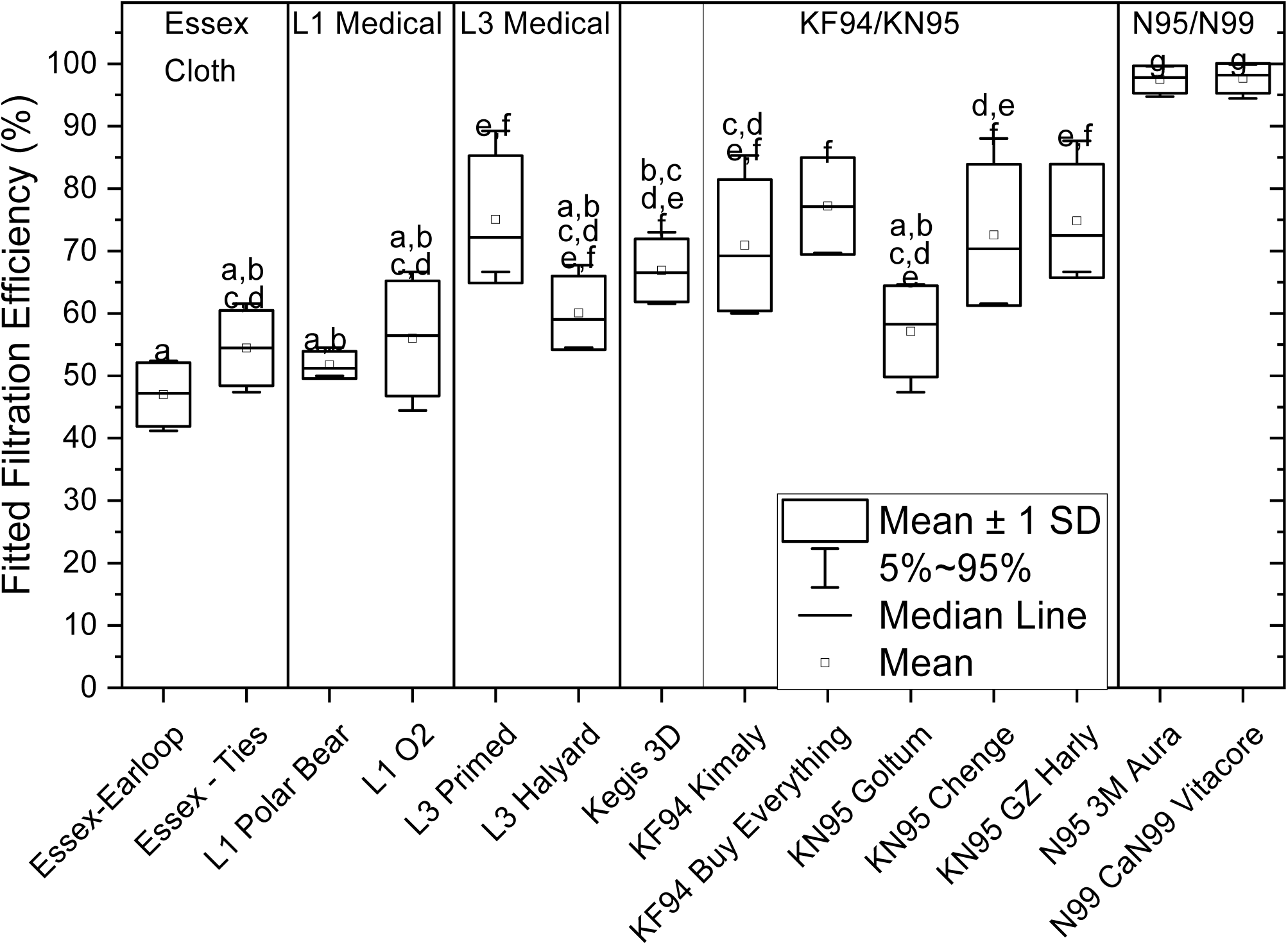
Fitted filtration efficiency for cloth (2), L1 (2) and L3 (2) certified medical masks, a non-certified Kegis 3D mask (purchased as a KN95 look-alike), KF94s (2), KN95s (3), and for respirators, N95 and CaN99. Footnote: N = 4. Bars present mean and standard deviation (SD) and whiskers showing 5 - 95 confidence values. Data were normal by Lillefor’s test. Letters above whiskers indicate statistical groupings according to Tukey’s post hoc comparisons. A shared letter for two mask types signifies no difference between those types; absence of a shared letter signifies a significant difference p<0.05.

### Mask hacks

Tested on 10 participants, median filtration efficiency of the level 1 and level 3 mask was not improved by wearing on ear guards, scrub cap or by knot-and-tuck (p <0.05, figure 2). Both masks were improved by braces designed to improve edge seal, reaching median filtration values of 85-90% for level 1 mask-and-brace combinations and 93–94% for level 3 mask-and-brace combinations (p <0.05).

**Figure 2.**
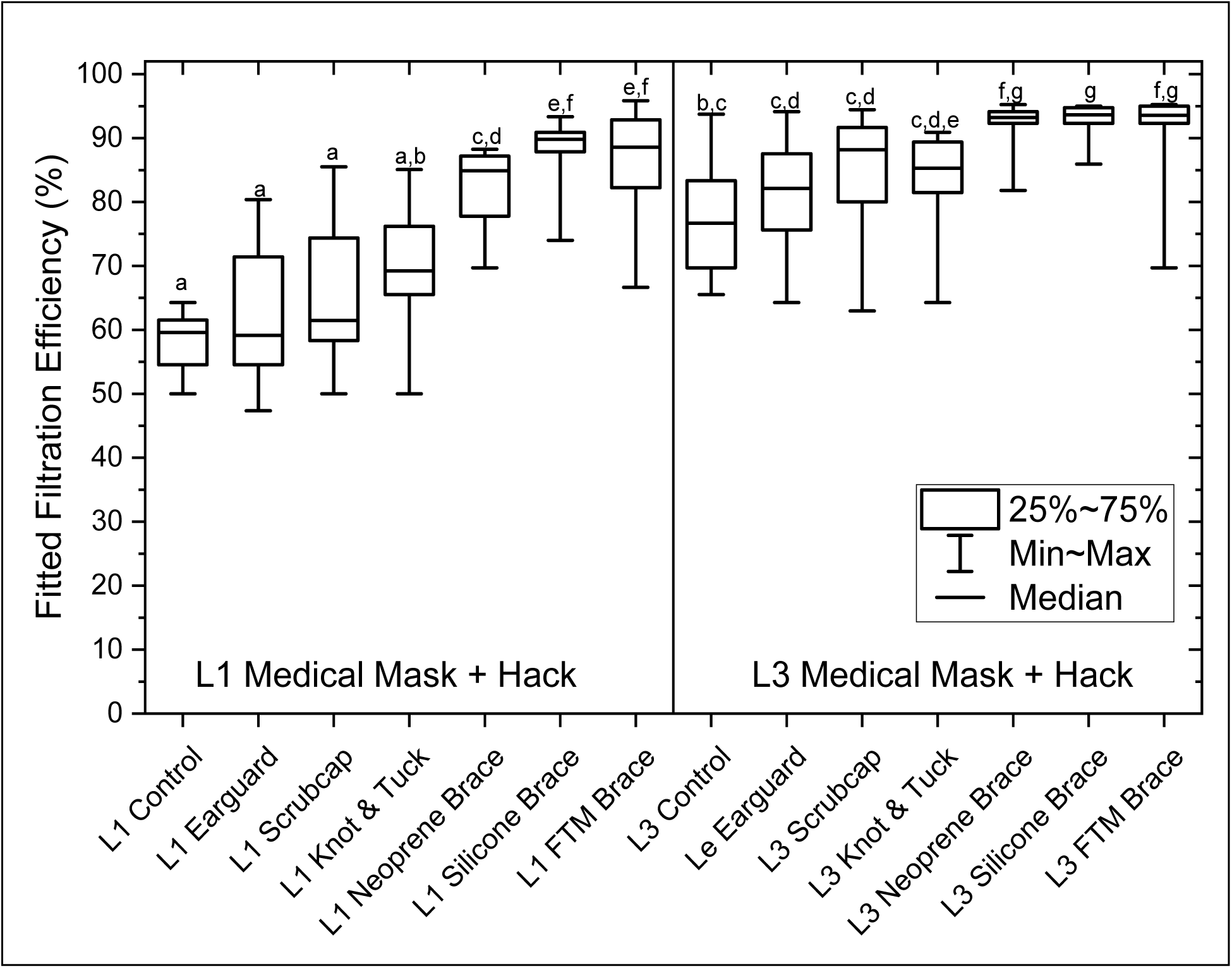
Box and whisker plot showing the effect of minor modifications, or hacks, to a certified level 1 Polar Bear mask and to a certified level 3 Halyard mask. 10 participants, 1 replicate. Footnote: N = 10. Data were not normal by Lillefor’s test. Kruskal-Wallis with Conover-Inman post hoc comparisons. Boxes show interquartile range and whiskers minimum and maximum. Letters denote groups which are statistically similar and dissimilar: a shared letter for two mask types signifies no difference between those types; absence of a shared letter signifies a significant difference p<0.05. Neoprene brace made using downloadable, public domain, template from Fix The Mask and recommended materials; silicone brace designed at McMaster University; FTM brace - proprietary Fix-The-Mask brace. L1 and L3 controls were retested on these participants as part of this panel; estimates differ slightly from those in figure 1.

### Overmasking

Tested on 6 participants (3 replicates per individual), mean filtration efficiency for the Essex mask on earloops was 47%, for the Essex on overhead ties 55%, and the level 1 mask 52%, p >0.05; the level 3 mask filtered 70%, p <0.05 (figure 3). Overmasking the Essex-on-earloops with another on earloops did not improve filtration but overmasking with an Essex-on-ties improved filtration to 66% (p <0.05). For both the level 1 and level 3 masks, overmasking with an Essex mask improved filtration. The level 1 mask overmasked with Essex-on-ties (84%) performed better than level-1–Essex-on-earloops (73%), both exceeding the level 1 mask control (all p < 0.05). The level 3 mask overmasked with Essex-on-earloops (83%) exceeded the control (p < 0.05) as did level-3–Essex-on-ties (92%; p <0.05), but the difference between the two was not statistically significant.

**Figure 3.**
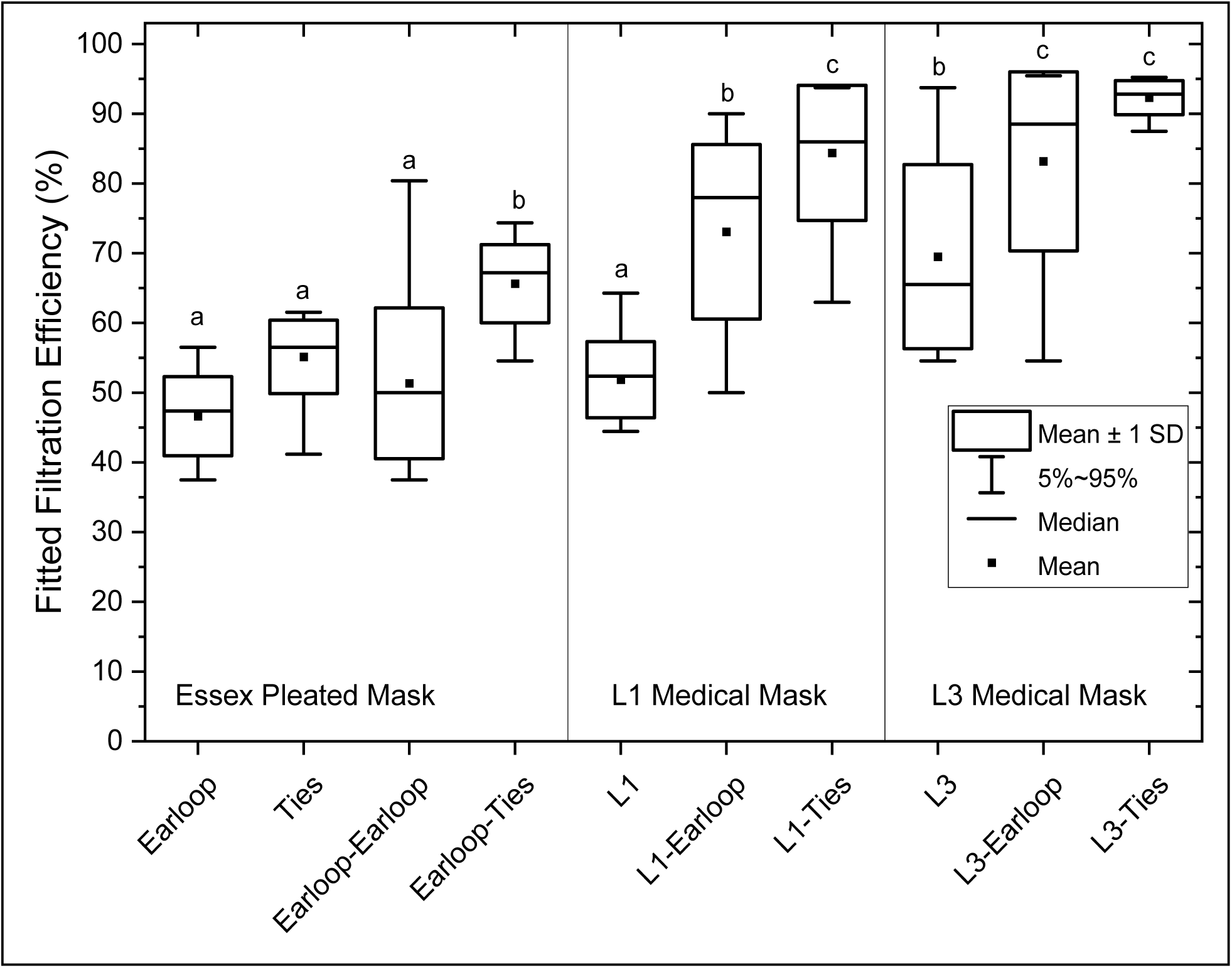
Effects of overmasking with Essex masks on earloops and on overhead ties on fitted filtration efficiency. The top graph shows (left) the Essex mask on earloops (Earloop) and on ties (Ties) worn alone, followed by Essex-on-earloop with a second Essex-on-earloop as an overmask (Earloop-Earloop), and by Essex-on-earloop with a second Essex mask on ties as an overmask (Earloop-Ties). The centre panel shows the level 1 certified Polar Bear mask worn alone (L1), with an Essex-on-earloop as an overmask (L1-Earloop), and with an Essex-on-ties as an overmask (L1-Ties). The right panel shows the level 3 certified Halyard mask worn alone (L3), with an Essex-on-earloop as an overmask (L3-Earloop), and with an Essex-on-ties as an overmask (L3-Ties). Footnote: N = 6, 3 replicates. Data were normal by Lillefor’s test. ANOVA with Tukey’s honestly significant difference for post hoc comparisons was used. Mean and median; boxes show one standard deviation (SD); whiskers show 95% confidence intervals. Letters denote groups which are statistically similar and dissimilar: a shared letter for two mask types signifies no difference between those types; absence of a shared letter signifies a difference. Earloop: Essex mask worn on elastic earloops. Ties: Essex mask worn on overhead cloth ties. L1 level 1; L3 level 3.

### Subjective assessment

Data on subjective assessment of leaks, glasses fogging, and discomfort are shown in the supplementary tables 2–3 and supplementary figures 3–5. Leak scores varied by mask type, but not with hacks or overmasking. Glasses fogging showed few statistically significant differences in any substudy. Discomfort scores ranged from 2 (comfortable) to 6 (uncomfortable) and differed across mask/respirator types. By category respirators and medical masks had better comfort than some KF94 and KN95s (p <0.05) but within in each category there were high- and low-comfort masks. Braces, knot-and-tuck and overmasking generated lower comfort compared with controls (all p <0.05). For braces this represented a trade-off between fitted filtration efficiency and comfort. Comfort scores were not related to fitted filtration efficiency.

Subjective assessment of mask leaks and glasses fogging were both significantly related to fitted filtration efficiency (p <0.05, figure 4). Leak assessment was the stronger predictor, explaining 22% of fitted filtration efficiency, whereas glasses fogging explained 4% of variation. Trials across masks with scores of 1 (no leaks detected) achieved mean filtration efficiencies of 85%, scores of 2 (imperfect seal to face) 75%, 3 (minor leaks) 72%, 4 (minor leaks in multiple areas) 65% and scores between 5-7 (major to severe leaks) averaged 59-60%.

**Figure 4.**
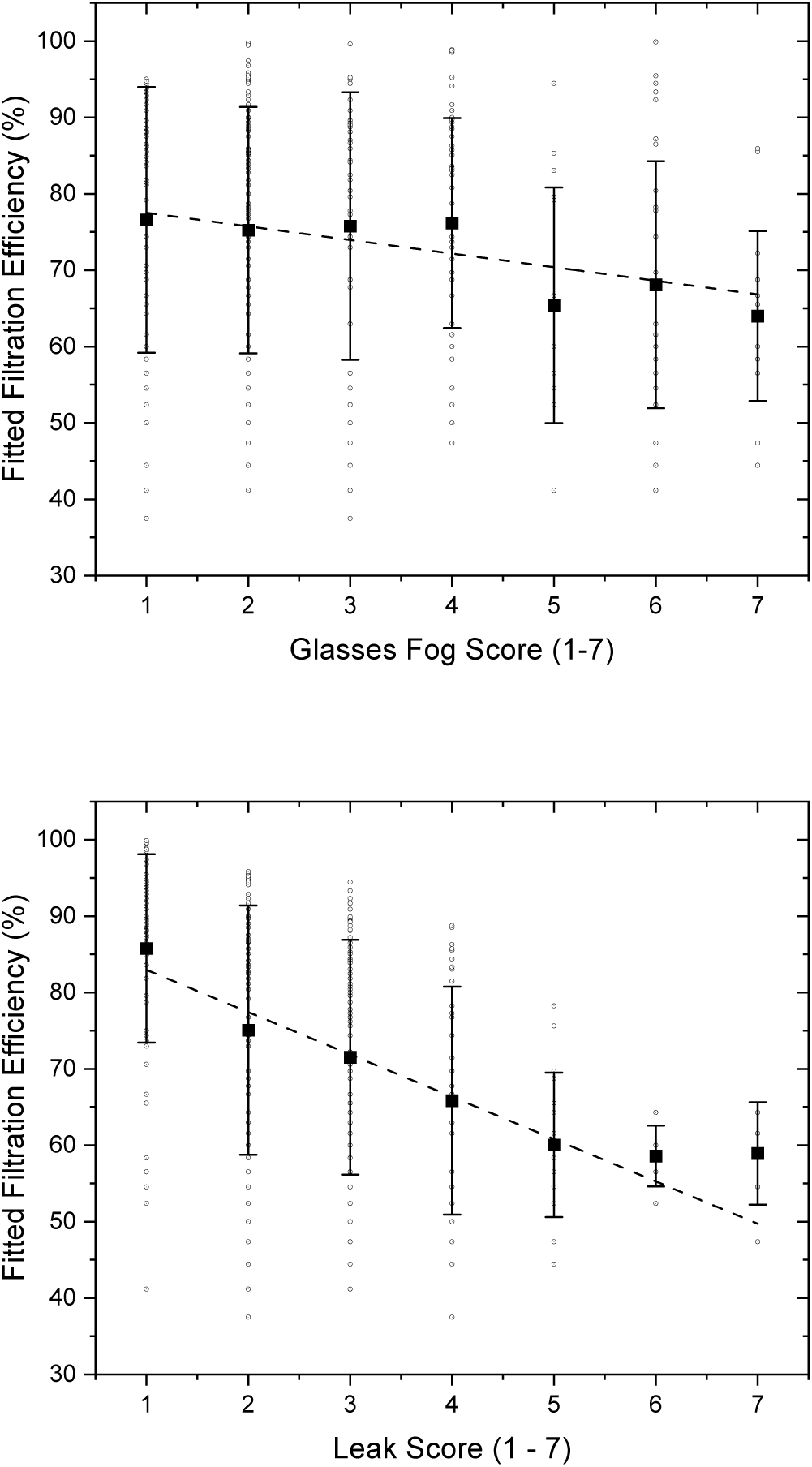
Relationship between fitted filtration efficiency and glasses fog, and between fitted filtration efficiency and subjective leak. Glasses fog and subjective leak were assessed before fitted filtration efficiency was measured. Top graphic fitted filtration efficiency against glasses fog score across data generated for each sub-study. Open circles are raw data, squares are means and whiskers are standard deviations for each score category. Dashed line is the linear regression fit: FFE = -1.78±0.48*Glasses Fog Score + 79.3±1.7; R^2^ = 0.04; p<0.001, df = 338. Bottom graphic presents data against leak score. Dashed line regression fit: FFE = -5.5±0.6*Leak Score + 88.5±1.7; R^2^=0.22; p<0.001, df =338.

### Facial measurements

There was no association between the facial distances bizygomatic distance and menton-sellion length, measured as if for clothing, with a piece of cord that traversed the bridge of the nose and the tip of the nose, respectively, and the same distance measured with calipers: R^2^ 0.03; p=0.53 and R^2^ 0.09; p=0.27, respectively (supplementary figure 6).

## Discussion

We found that well-fitting cloth masks on earloops and ties filtered at 47-55%; this was comparable with level 1 certified masks (51-60%; infographic, supplementary figure 7). This is not widely known: the filtration properties of the materials are very different.^29^ The better edge seal of the well-designed cloth mask makes up for the relatively poor filtration of woven cotton;^7, 29^ the excellent filtration (>95%) of the material used in level 1 masks^27, 29^ is let down by poor fit and edge leak. Level 3 masks were variable (60-77%).

In the media,^40^ in CDC messaging,^41^ and in epidemiologic research,^13^ KF94/KN95-type masks are often conflated with N95s, FFP3s, and CaN99s. We observed 57-77% filtration for the KF94/KN95s. The previous literature is limited to two masks, one with 55% filtration efficiency (7 participants, 1 replicate),^42^ and the other 84% (≥8 participants).^43^ We found most, but not all, KN95/KF94s performed better than cloth and level 1 masks, similar to level 3 masks, but not as well as the N95 and CaN99. The material in KF94/KN95s is an excellent filter; the Chengde and the GZHarley masks were tested by NIOSH and passed, with all masks providing greater than 95% filtration when tested with the edges glued to a flat plate in keeping with NIOSH Standard Test Procedure (STP) TEB-APR-STP-0059.^44^ The difference between them when worn must result from design and fit, including the type of head attachment: N95s and CaN99s have overhead elastic.

In contrast, we found 97-98% filtration for N95 and CaN99 respirators that were not formally fit-tested: this is close to the fit-testing threshold of ≥99% (ie, fit factor ≥100). Other studies of non-fit-tested respirators have reported 82% to 98% filtration.^42, 45–49^ Both our respirators were of the novel 3D design, which may fit a wider variety of faces than the cup designs in common use before the pandemic. In terms of the fraction reaching the participant, the respirators let through up to 3% of particles and the KF94/95s up to 43%: a 14-fold difference in exposure. We argue that this difference should be more widely known and that, provided PPE supply for frontline workers is secure, N95s and CaN99 should be recommended for community use.

Previous studies of fitted filtration efficiency for medical and non-medical masks have been limited by very small sample sizes and incomplete descriptions of the masks. For cloth masks, our data (47-55%) are in keeping with the higher end of reported fitted filtration efficiencies: 27% (3-ply cotton on earloops, 0.02-3µm particles, 1 participant, 4 replicates)^5^; 28% (2-ply, 3-ply and 4-ply polyester, cotton and poly-cotton masks on earloops, 0.1µm particles; 3-4 participants, 1 replicate)^50^; 50% (2-ply cotton T-shirt fabric, on overhead elastic ties, <0.1µm particles, 21 participants, 1 replicate)^51–53^; 52% (head attachments and material not reported, 5 designs including 1-ply and bandana fabric, <0.1µm particles, 3 participants, 1 replicate).^42^ This likely reflects the carefully-designed Essex mask, in contrast with random variation from haphazard sampling.^5, 42, 50^

Level 1 masks were associated with filtration of 52-60%, and level 3 masks with filtration of 60-77%. Previous studies have included a single mask, often incompletely specified, and with a limited number of observations: 39% (procedure mask on earloops, 0.02-3µm particles, 1 participant, 4 replicates)^5^; 72% (surgical mask with ties, 0.02-3µm particles, 1 participant, 4 replicates)^5^; 69% (unknown design, 0.1µm particles, 7 participants, 1 replicate)^42^; 80% (level 1 equivalent on ties, 0.1µm particles, 21 participants).^54^ The difference in fitted filtration we observed between some level 3 and level 1 masks has not previously been reported and likely results from better fit: the difference in material flat filtration for these two standards is small (98% v 95% for 0.1µm latex).^27, 28^ The level 3 masks we studied were larger top-to-bottom than the level 1 masks, had foam at the nosepiece, were softer, and appeared to conform better to participants’ faces.

Ours is the largest study to date to examine a comprehensive range of minor modifications (mask hacks), and overmasking; and the first to examine the effects on both level 1 and level 3 masks. With a larger sample size than any previous study (10 participants), we were not able to confirm the improvements seen for earguard and knot-and-tuck in previous studies (1 participant, 4 replicates^5^; 3-4 participants^50^), though these methods may increase fitted filtration for some individuals. We confirmed the findings (1 participant, 4 replicates^5^; 11 participants^55^ and 3-4 participants^50^) that external braces greatly improve the efficiency of certified masks (infographic, supplementary figure 7). We have previously shown that braces have little effect over cloth masks.^56^

We found that overmasking with a carefully-designed cloth mask was associated with increases in filtration efficiency. Our study of 6 participants with 3 replicates is a significant addition to previous studies (1 participant, 4 replicates, using nylon hosiery^5^; 3-4 participants, overmasking^50^) which also found overmasking effective. Overmasking should be encouraged when N95s are not available.

Our work provides the first direct comparison of earloops and ties, finding ties superior when used as an overmask with a level 1 mask, and a non-significant tendency to superiority when worn alone or over a level 3 mask. This is in keeping with the idea that though earloops are convenient for donning and doffing, overhead attachments produce a closer fit and less edge leak, and with the observed differences between KF94/KN95s which have earloops, and N95/CaN99s which have overhead elastic.

Our KN95/KF94 sampling strategy simulated an informed non-expert purchaser: we recognize that our sample is small.^57^ It is a limitation of our work that we did not include non-certified pleated disposable masks or CaN95s.

We recognize additional limitations (supplementary material, p 36). We studied one cloth mask, a design that was created and refined by a panel of community experts, optimized, and produced at scale (55,000 masks) by non-expert sewists for distribution to marginalized communities. The ASTM standard for barrier face coverings, F3502, which is a source-control standard, not designed to assess the degree of protection for the wearer, defines filtration efficiency of a mask edge-sealed to a plate, not worn^58^; we did not perform this test. Finally, we examined masks through the lens of wearer protection; we provide no data on source control, for which there is a wealth of data^59^ but no standard methodology.^60^ However, previous studies suggest a relationship between the two,^45, 50, 61^ and F3502 recognizes Portacount testing as the only available quantitative standard for assessing fit for source control.^60^

Strengths of our study are the large number of participants compared with previous studies, the characterization of the participants, the characterization of the masks, and the inclusion of a variety of different masks, including KN95/KF94s, and variety of mask hacks, in the same study. We used human participants and studied 0.02 to 1 µm particles, which is the smaller end of the particle size range that is thought to be most clinically relevant.^62–66^

## Conclusions

Well-designed cloth masks exhibited fitted filtration for submicron aerosols similar to that of level 1 masks: around 50%. Level 3 masks and KN95/KF94s performed around 70%. Overmasking of certified level 1 and 3 masks with cloth masks was effective and external braces over level 1 and 3 masks were highly effective in improving filtration efficiency. Filtration at or close to occupational health standards for PPE was observed only for N95s and CaN99s. Increased clarity on the relative aerosol filtration properties of masks other than respirators will be useful to public messaging about masks and in the interpretation of implementation studies.

## Data Availability

All data are summarised in the manuscript and supplementary figures.

## Acknowledgements

The Centre of Excellence in Protective Equipment and Materials and McMaster University are built on the traditional territory of the Haudenosaunee and Anishinaabe first nations, recognized in the 1701 Dish with One Spoon Wampum.

We thank Bryan Herechuk, Jeff Mallany, Anna Dorey, Seema Sharma, Hamilton Health Sciences, Hamilton, St Joseph’s Healthcare Hamilton, Hamilton, ON for mask donations, materials, advice and encouragement.

Windsor Essex Sewing Force (WESF) provided cloth masks, advice and encouragement. They place their pattern in the public domain with this publication.^30, 31^

We thank the stakeholder group Cloth Mask Knowledge Exchange (CMKE) for advice and encouragement at every stage of this work.

We dedicate this work to the memory of our CMKE colleague Helen Brunet.

